# Regional Anesthesia in Intensive Care: An Overview in Tunisia

**DOI:** 10.1101/2024.06.22.24308393

**Authors:** Dorra Nouri, Fatma Ezzahra Nouri, Aicha Rabai

## Abstract

Regional anesthesia (RA) is increasingly used in intensive care in Tunisia, but challenges persist to ensure optimal practice. We conducted a multicenter study involving post-operative and polyvalent intensive care units, both private and public. Resident physicians(44.7%) are sensitized to quality of care, but gaps remain. Attending physicians (63.3%) often report the absence of pain management committees (PMCs) and written protocols for RA. The majority express a need for continuous training, particularly on RA. High-fidelity simulation is the preferred format for learning. RA is commonly used in intensive care (97.2%), mainly epidural (76.4%) and femoral nerve blocks (54.9%). Ultrasound is widely preferred for guiding procedures (77.5%). The main areas of RA application are thoracic (94.4%) and limb trauma (64.8%). The ANI is the preferred pain monitoring tool (49.3%). Improving training and infrastructure is necessary for optimal RA practice in intensive care in Tunisia.

## Introduction

Locoregional anesthesia plays a crucial role in the perioperative management of patients. Compared with systemic administration of hypnotic or anesthetic agents, this anesthetic technique has been shown to reduce perioperative morbidity and mortality [1-4]. Although this practice is becoming increasingly common in operating rooms, thanks in particular to ultrasound guidance, its use in the intensive care unit remains limited [1], despite the high incidence of pain in intensive care units. Indeed, this is estimated at 30-50% of patients at rest, and up to 80% of patients when certain invasive procedures are required for their care [5]. As an anesthesiologist, it is important to take stock of the use of this technique, in our surgical and general intensive care units. In this article, we will examine current data on the use of locoregional anesthesia in intensive care units in Tunisia, in order to identify the factors explaining the disparity in its use, and the challenges to be met for optimal practice of this technique.

## Methodology

This was a prospective, observational, descriptive, cross-sectional, and multicenter survey. The survey was based on a self-administered questionnaire assessing the use of regional anesthesia in intensive care units in Tunisia. The questionnaire was intended for anesthesiologists and intensivists, whether resident or attending physicians, working in a surgical or general intensive care unit, whether in a private or state institution. The survey was conducted over a period of 4 months from August 1st to November 30th, 2023. The anonymized questionnaire consisted of 33 questions. The information collected through a link provided by the Google Forms software included demographic data such as gender, status, type of institution, and intensive care unit. In addition, medical data were collected, relating to the practice of regional anesthesia (incidence of pain, indications, frequency of use, reasons for not using, type of block and method used, monitoring, access to intralipids, etc.). Finally, our questionnaire aimed to determine the level of continuing education in the different intensive care units and the specific training expected in this field. Statistical analysis was performed using SPSS software. For qualitative variables, results were described in terms of frequency and percentage. For the comparison of means, we used the Chi-square test with a significance level set at 0.05. Percentage values are given with a 95% confidence interval.

## Results

### Demographic Data of Participants

At the end of our survey, we collected responses from 71 anesthesiologists and intensivists. The main demographic characteristics are presented in Table I. The majority of respondents were female, with a sex ratio of 0.73. Regarding the institution, the majority of participants worked in public institutions (81.7%), while 18.3% worked in the private sector. The survey was conducted in 48% of cases in general intensive care units and in 23% of cases in postoperative intensive care units. Responses were collected from resident physicians in 53.3% of cases, with their distribution represented in Table I.

**Table I:**
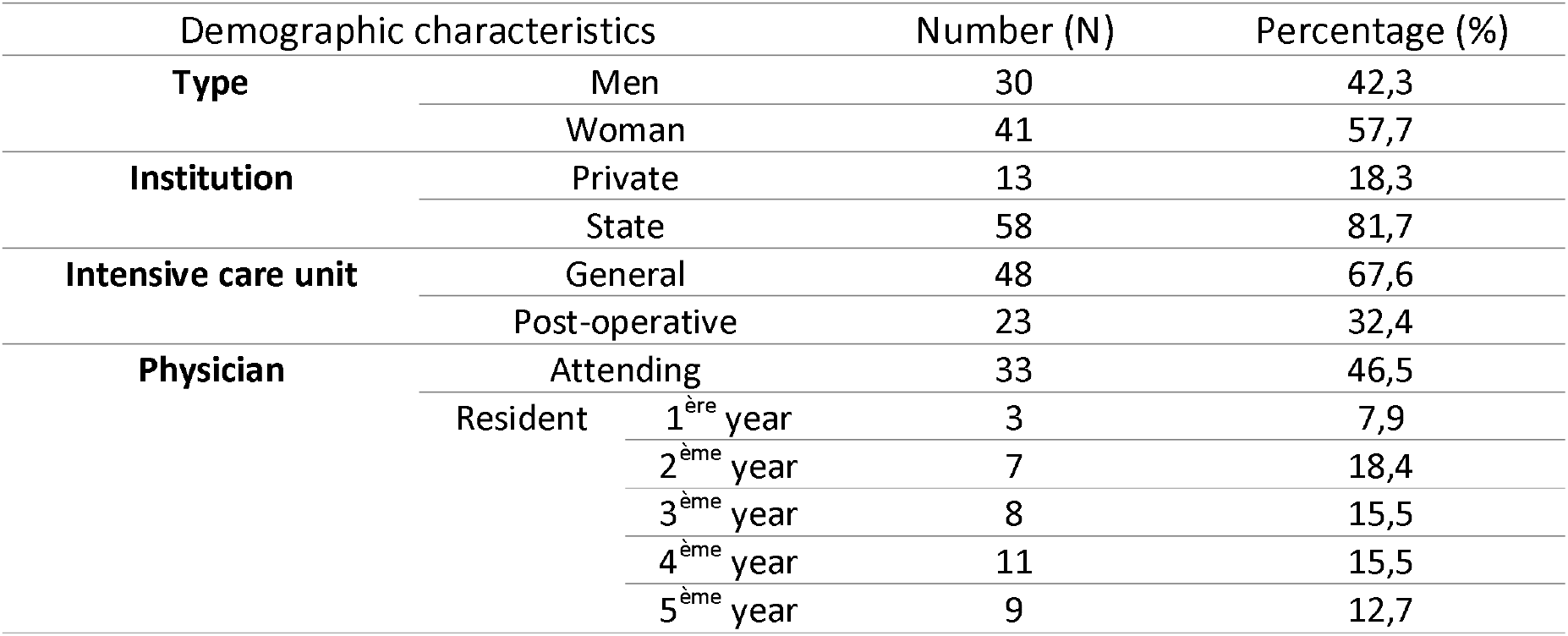
Demographic characteristics of surveyed physicians.

### Practice of Regional Anesthesia

The assessment of pain levels in the ICU by the participants is summarized in the following figure (Figure 1):

**Figure 1:**
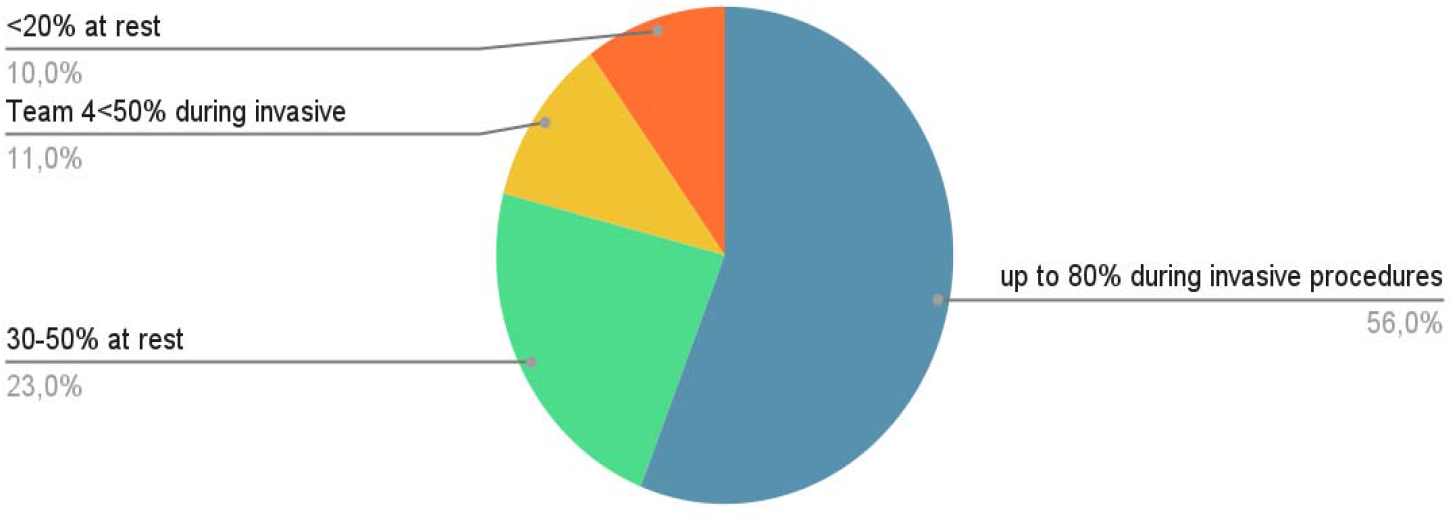
Pain Assessment in Intensive Care by Participants.

For suitable candidates for the practice of regional anesthesia in intensive care, the respondents’ answers are summarized in the following figure (Figure2):

**Figure 2:**
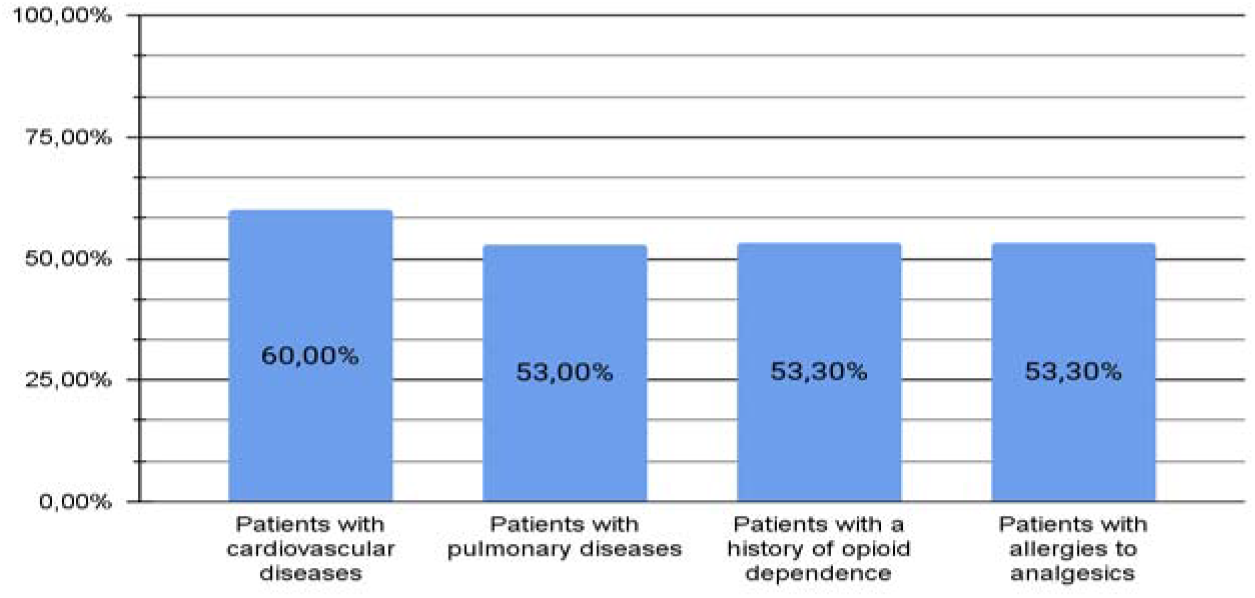
Suitable candidates for regional anesthesia practice in intensive care.

In clinical practice, the indications for regional anesthesia in intensive care as reported by the participants are presented in the following figure (Figure 3):

**Figure 3:**
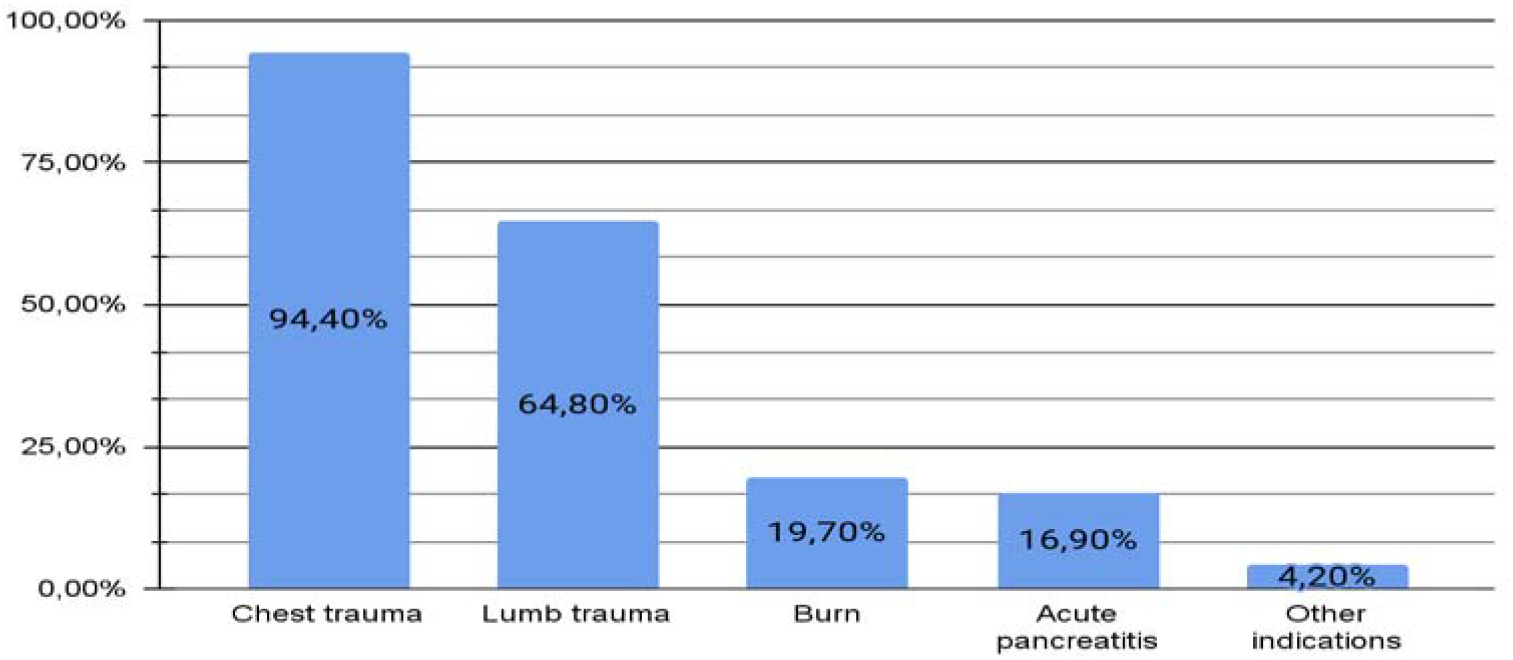
Indications for Regional Anesthesia in Intensive Care.

Regarding the use of regional anesthesia in intensive care, the majority of investigators confirmed its use in this practice, with a rate of 97.2%. It is also relevant to assess the frequency of this practice among respondents. Our results indicate that this technique was used commonly or occasionally in 74.3% of cases. Regarding the types of blocks commonly used, epidural anesthesia was the most frequent, used in 74.6% of cases, followed by femoral nerve block, used in 54.9% of cases. The main blocks used are summarized in the following table (Table II):

**Table II:** The main blocks used in intensive care.

| Block type | Frequency (N) | Percentage (%) |
| --- | --- | --- |
| Peridural | 53 | 74,6 |
| Femoral block | 39 | 54,9 |
| Iliofacial block | 28 | 39,4 |
| Paravertebral block | 21 | 29,6 |
| Axillary block | 4 | 5,6 |
| TAP block | 4 | 5,6 |

Ultrasound guidance was the most frequently used technique for performing regional anesthesia in intensive care, with a rate of 77.5%. The different methods used are represented in the following figure (Figure 4):

**Figure 4:**
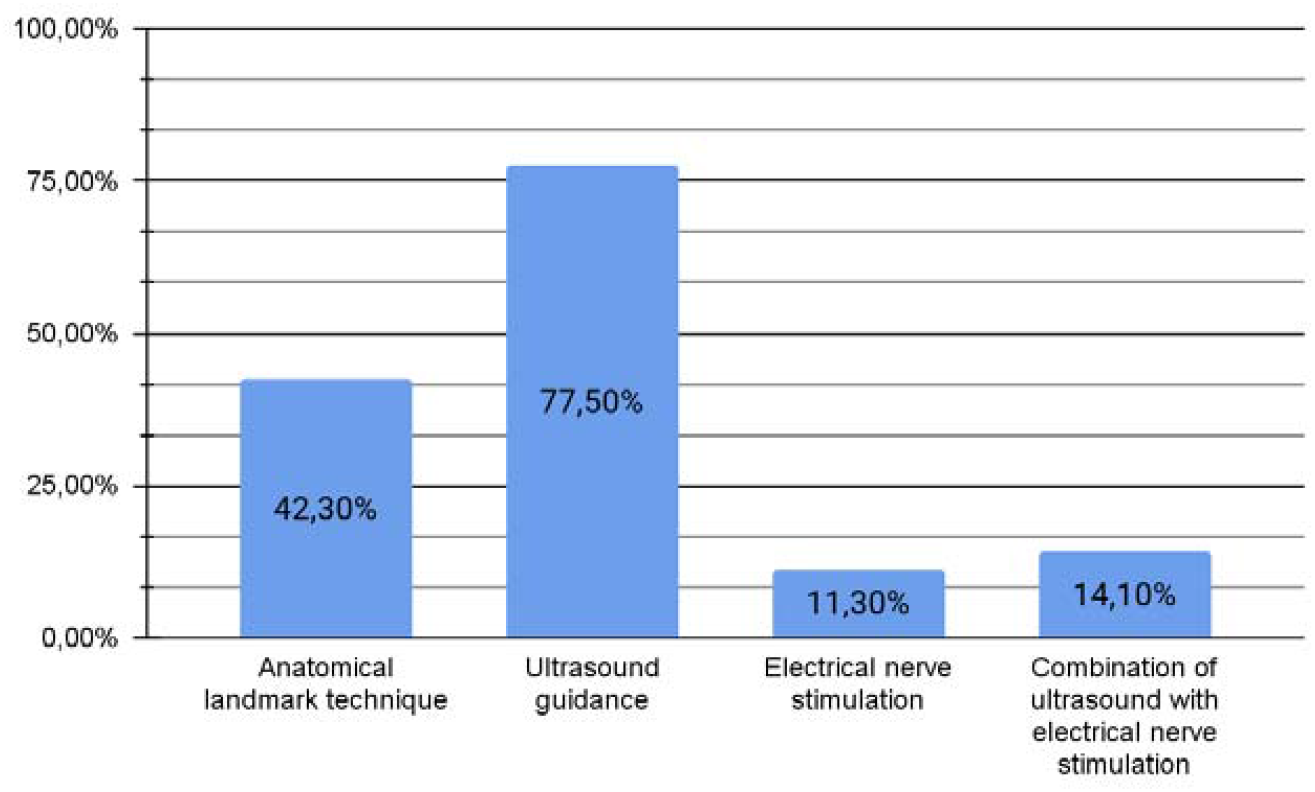
Different methods used in performing regional anesthesia in intensive care.

Only 2.8% of participants have never used regional anesthesia in intensive care. Among the reasons cited by these participants are: technical problems such as unavailability of ultrasound, regional anesthesia needles, etc., lack of knowledge of anesthesia techniques, and lack of training for team physicians.

Our investigation also focused on the availability of intralipids in the various intensive care units. Indeed, our investigators reported easy accessibility in 49.3% of cases. Thus, the unavailability of intralipids was observed in 50.7% of cases.

Regarding pain monitoring in intensive care, the Analgesia Nociception Index (ANI) was the most chosen method by respondents, with a rate of 49.3% (Figure 5):

**Figure 5:**
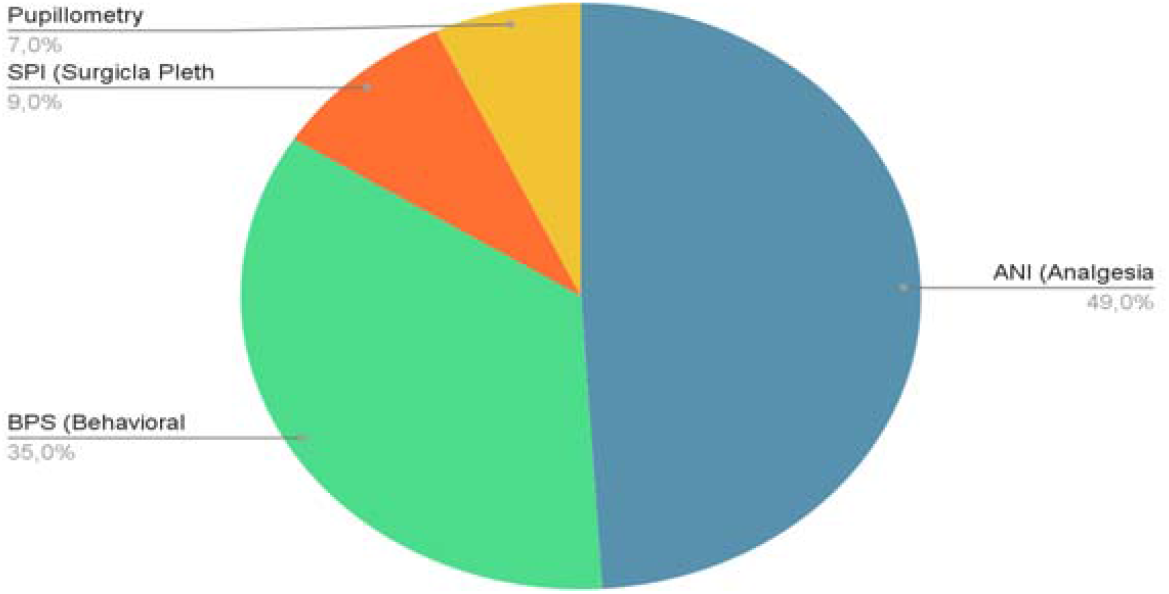
Pain monitoring methods used in intensive care.

Regarding the evaluation of the effectiveness of regional anesthesia in intensive care, the majority of investigators used the Visual Analog Scale (VAS) in 81.7% of cases. This was followed by a decrease in the need for opioid analgesics in 43.7% of cases, and finally the Analgesia Nociception Index (ANI) in 32.4% of cases.

### Training in Regional Anesthesia

Results regarding the level of training of resident physicians revealed that 44.7% of them were sensitized to concepts of quality care, procedures, protocols, or audit missions. The majority expressed an interest in training focused on quality care and risk management (92.1% of cases). All resident physicians wished to participate in training on regional anesthesia in intensive care. As for the preferred learning format, 60.5% of them opted for high-fidelity simulation. Other preferred formats included procedural simulation (52.6%), tutorials (44.7%), bedside teaching (36.8%), staff room presentations (28.9%), reading (5.3%), and summary posters (5.3%).

For attending physicians, 63.3% of them reported the absence of a Pain Control Committee (PCC) in their institutions and the absence of written procedures describing the conduct of regional anesthesia in 57.6% of cases. In cases where these were present, investigators indicated their permanent presence in the intensive care units, in 92.3% of cases. These procedures were the subject of staff training in 61.5% of cases and audit missions in 46.1% of cases. Furthermore, 90.9% of attending physicians wished to participate in training on regional anesthesia in intensive care, whether in the form of tutorials, high-fidelity simulation, or procedural simulation (in 21.2% of cases), reading, staff room presentations, or immersion days (in 18.2% of cases), or summary posters or bedside teaching (in 15.2% of cases).

## Discussion

Our study has shown a notable increase in the use of locoregional anesthesia (LRA) in Tunisian intensive care units (ICUs) in recent years. The uniqueness of our study lies in its multicenter nature, involving postoperative and general ICUs, both in the private and public sectors. Moreover, to our knowledge, this is the only study in Tunisia and worldwide focusing on this topic. However, the relatively small number of respondents is a limitation.

In addition to its proven effects in the literature for reducing perioperative morbidity and mortality [1-4], LRA is considered an effective analgesic strategy in many post-traumatic or postoperative clinical situations in the ICU. Regarding the estimation of pain levels in intensive care units by the participants, 56% of them consider this level to reach up to 80% during invasive procedures. This value was adopted by Yoanna Skrobik and her team in their publication of clinical practice guidelines for the prevention and management of pain in ICU patients in 2013 [5]. Other studies have specified the clinical repercussions of inadequately relieved pain on ICU patients, such as activation of the sympathetic nervous system, myocardial ischemia due to imbalance in myocardial energy balance, lipolysis and muscle wasting, reflex ileus, immunosuppression, and cognitive disorders [6]. Thus, pain management is an essential element to ensure patient comfort and limit suffering.

For the selection of the target population, our respondents consider patients with cardiovascular diseases as the most appropriate candidates to benefit from this technique, in 60% of cases. Next are patients with a history of opioid dependence and patients with an allergy to local anesthetics in 53.5% of cases, and finally, patients with pulmonary diseases in 53% of cases. As described above, several studies emphasize the role of LRA in pain prevention, and therefore in the cardioprotection of vulnerable patients. Other studies have demonstrated the effect of LRA, particularly epidural anesthesia, in reducing the risk of atrial fibrillation and deep vein thrombosis [2,7]. The effect on respiratory function was also highlighted by several studies, especially in patients with chronic obstructive pulmonary disease. Indeed, in combination with general anesthesia, LRA, and particularly epidural anesthesia, was associated with a reduction in the risk of pneumonia, dependence on mechanical ventilation, and re-intubation [8]. We also highlight the important role of local anesthetics in morphine-sparing and reducing the use of opioids. The latter have several deleterious effects such as respiratory depression, urinary retention, and issues with dependence and withdrawal [1]. All these data underscore the crucial role of LRA in the management of fragile patients in intensive care.

Regarding the fields of application of LRA in intensive care, thoracic trauma constitutes the most frequent indication for the participants. Indeed, 94.4% of respondents opt for LRA as an effective analgesic strategy against pain and the onset of respiratory complications (such as atelectasis or post-traumatic pneumonia) [1]. In the case of unilateral lesions, the paravertebral block is preferred, while the epidural is proposed for managing complex or bilateral lesions [9]. Limb trauma is also another field of LRA implementation in intensive care, chosen by 64.8% of participants. Indeed, peripheral blocks provide effective analgesia while avoiding the harmful effects of systemic analgesia on major functions [10]. In this context, several studies recommend the use of a perineural catheter to prolong the duration of the analgesic block [11] and promote the terminal vascularization of traumatized segments thanks to the sympathetic block induced by the peripheral block [12]. According to our study, LRA also finds its place in the management of burns in intensive care, in 19.4% of cases. In this context, the team of Chaibdraa et al. performed 634 LRA procedures on burn patients, primarily targeting the lower limbs (75% of cases, mainly by femoral block) over a period of 3 years. The efficacy was 95% under ultrasound guidance, with a relatively low complication rate (3%) and no deleterious effects [13]. However, the use of LRA in these patients can be challenging: risk of infection, increased absorption of local anesthetics due to the inflammatory spread (less efficacy and increased toxic risk) [1]. According to the results of our study, LRA can also be a useful option for the management of acute pancreatitis in intensive care, in 16.9% of cases. In this context, several studies have emphasized the importance of LRA, particularly epidural anesthesia, in pain relief and improving splanchnic perfusion without increasing the septic risk in these patients [14–18].

In practical terms, the use of LRA in intensive care in Tunisia is widespread. A substantial 97.2% of our respondents frequently use this analgesic strategy. The rapid learning curve and the availability of the necessary technical facilities explain the easy access to this practice in various intensive care units. Among all the blocks described, the epidural block is the most commonly used analgesic block, performed by 76.4% of our respondents, followed by the femoral block, used in 54.9% of cases. This can be attributed to the frequency of ICU admissions for polytrauma patients (with thoracic and peripheral components involving the lower limbs) and the postoperative management of elderly patients undergoing surgery for upper femur fractures or major abdominal surgery (in the context of cancer treatment). The ease of learning these two analgesic blocks may be another explanation.

Ultrasound is the reference technique for performing LRA in intensive care, adopted by 77.5% of our respondents. This technique, recommended by various scientific societies, has secured these procedures in operating rooms and ICUs: it increases the success rate of the block while limiting the risk of local anesthetic toxicity by reducing the required volumes and concentrations of anesthetic agents [10].

However, a small proportion of our respondents did not use LRA in their clinical practice in intensive care (2.8% of participants). Among the challenges faced by these healthcare professionals, the unavailability of necessary technical equipment (ultrasound machines, LRA needles, etc.) was cited. The lack of training and unfamiliarity with the anesthetic technique also posed obstacles for several physicians. Consequently, it seems essential to generalize access to the necessary resources and training in this field to ensure equal opportunities among different anesthesia and intensive care teams in Tunisia. Another constraint reported by the respondents was the unavailability of intralipids in intensive care units, observed in 50.7% of cases. As part of the initial checklist before performing each block, the availability of intravenous lipid emulsions ensures the safety of performing LRA in intensive care [19]. Therefore, it is crucial to ensure the accessibility of intralipids to guarantee the safety of this analgesic strategy. Nevertheless, we cannot compare our data with those in the literature. To our knowledge, no published national or international study has addressed the practice of regional anesthesia in intensive care.

To assess the level of pain in intensive care, 49.3% of our respondents consider the ANI (Analgesia Nociception Index) to be the most suitable monitoring method, followed by the BPS (Behavioral Pain Scale) in 35% of cases. Derived from heart rate variability, the ANI is used in various studies to measure the response to nociceptive stimuli during surgery, thus contributing to the optimization of analgesic administration in anesthetized patients [20,21]. The BPS (Behavioral Pain Scale) is recommended for monitoring pain in unconscious patients or those unable to communicate, in line with the common consensus on sedation-analgesia in intensive care by the French Society of Anesthesia and Intensive Care (SFAR) and the French-speaking Intensive Care Society, published in 2008 [22]. However, concerning the evaluation of LRA effectiveness in an anesthetized patient, our participants preferred the Visual Analog Scale in 81.7% of cases due to its simplicity and reproducibility. Nonetheless, it is important to note that this scale requires a degree of alertness and cooperation, which may be lacking in an intensive care patient. In this context, recommendations suggest using the pupillary dilation index to monitor the effectiveness of the analgesic block [22]. Therefore, given the variability of the adopted scales, implementing a standardized protocol tailored to each intensive care unit is recommended. The results related to the training level of residents reveal that awareness of care quality concepts, protocols, procedures, and the audit mission has not been observed in 55.3% of them.

However, a marked interest in training focused on care quality and risk management was expressed by the majority of our respondents (92.1%). The demand for training in regional anesthesia in intensive care was mentioned by all resident physicians, reflecting the recognition of the importance of this analgesic strategy in intensive care. The preferred learning format, with 60.5% opting for high-fidelity simulation, indicates a preference for interactive and practical methods. Other formats, such as procedural simulation, tutorials, and briefings in the staff room or at the bedside, demonstrate the variability of approaches desired by the resident physicians. As for the attending physicians, the results highlight significant gaps, notably the absence, in most cases, of a pain management committee (CLUD) and the lack of written provisions detailing the conduct of regional anesthesia in intensive care in more than half of the cases. However, these provisions are generally present in intensive care units (92.3%) whenever they exist. The training of nursing staff and audit missions related to these provisions reflect a commitment to ensuring better implementation of regional anesthesia in intensive care. In terms of continuing education, most attending physicians expressed their desire to participate in training programs on regional anesthesia in intensive care. Thus, developing targeted training programs that meet the specific needs of intensive care professionals seems essential to guarantee better implementation of regional anesthesia in intensive care.

## Conclusion

In summary, this study highlights the growing importance of regional anesthesia in intensive care in Tunisia while emphasizing the issues that need to be addressed to ensure optimal practice. To meet the increasing needs of healthcare professionals and ensure effective pain management in intensive care in Tunisia, additional efforts in continuing education, standardization of practices, and improvement of infrastructure are necessary.

## Data Availability

All data produced in the present study are available upon reasonable request to the authors

## Notes

**Disclosures:** We declare no conflict of interest.

### Competing Interest Statement

The authors have declared no competing interest.

### Funding Statement

This study did not receive any funding

